# A dual-mode targeted Nanopore sequencing assay for comprehensive *SMN1* and *SMN2* variant analysis

**DOI:** 10.1101/2024.02.22.24303180

**Authors:** Brad Hall, Sawsan Alyafei, Sathishkumar Ramaswamy, Shruti Sinha, Maha El Naofal, Fatma Rabea, Bryan J. Killinger, Gary J. Latham, Ahmad Abou Tayoun

## Abstract

**Background:** Spinal Muscular Atrophy (SMA) is one of the most common recessive disorders for which several life-saving treatment options are currently available. It is essential to establish universal SMA screening and diagnostic programs using scalable, cost-effective and accessible platforms to accurately identify all variation types, which is complicated by homologous *SMN1* and *SMN2* genes.

**Methods:** We developed a dual-mode PCR-based target enrichment that generates 2.7 to 11.2 kb amplicons spanning *SMN1* and *SMN2* genes for any-length nanopore sequencing. We trained a variant calling model that utilizes paralog-specific sequences and read-depth data to accurately detect sequence and copy number variants specific to each gene.

**Results:** We present results from the development, optimization, and external evaluation of this assay using over 750 samples, including cell lines, residual presumed normal blood donors, and patients with known *SMN1* and *SMN2* genotypes. The assay detects SNVs, indels, and CNVs with >98% accuracy across all sample sets, with a highly dynamic throughput range, relatively fast turnaround time, and limited hands-on-time. Together with the modest capital investment and consumable costs per sample, this assay can help increase access to SMA testing in low- and middle-income settings.

**Conclusion:** We describe a PCR/Nanopore sequencing assay and a customized analysis pipeline for the comprehensive and accurate detection of variation at the SMA locus and demonstrate its scalability, cost-effectiveness, and potential for the universal implementation of SMA screening and diagnostic programs.

**Human Genes:** *SMN1* survival of motor neuron 1, telomeric HGNC:11117 *SMN2* survival of motor neuron 2, centromeric HGNC:11118 *CFTR* CF transmembrane conductance regulator HGNC:1884

## Introduction

Spinal Muscular Atrophy (SMA) is the second most common autosomal recessive disease and the most common genetic cause of infant death (1). In around 95% of cases, SMA is caused by a homozygous deletion of the *SMN1* gene, which encodes for the survival motor neuron 1 protein (2, 3, 4); loss of this protein leads to degeneration of the motor neurons in the spinal cord and progressive muscle weakness, paralysis and, if untreated, premature death (5). Additional intragenic and structural variants have also been identified (6).

Both *SMN1* and its paralog, *SMN2* encode the same protein, SMN, and copy numbers of each within the genome can range from zero to four or more. SMA disease is modified by copy number status of the *SMN2* gene, which has >99.9% sequence identity to *SMN1* (7), though it does not produce a functional product because of a single nucleotide change in exon 7 that causes exon skipping and produces a truncated, nonfunctional protein (8). However, due to leaky expression of the full-length protein from the *SMN2* locus, its copy number status is inversely correlated with SMA clinical phenotype and disease severity (9, 8, 10). Patients at the severe end of the spectrum (SMA type I) often have one copy of *SMN2*, while those with a milder phenotype (SMA type IV) have greater than or equal to 4 copies of this gene (9, 11, 5).

Given its prevalence, life-threatening outcomes and, most recently, the availability of three life-saving medications approved by the US FDA (Zolgensma, Spinraza, and Evyrsdi), genetic diagnostic and screening programs have become essential for risk assessment, early detection, and timely patient treatment (12, 13). Such programs, however, require rapid, accurate, and comprehensive testing platforms to unambiguously detect and resolve single nucleotide (SNVs) and copy number variants (CNVs) in the *SMN1* and *SMN2* genes, as well as complex rearrangements leading to gene conversions (14) and silent carriers (15).

SMA testing strategies should also be highly scalable, cost-effective, and not require significant capital investment or complex infrastructure to support the deployment of universal screening programs in low-resource settings (16). These qualities are not only essential for equitable global access to screening but also important for characterizing disease epidemiology (incidence, prevalence and carrier frequencies) across populations. Such information can then inform the most appropriate screening and prevention strategies (17).

Since *SMN2* has high homology to *SMN1*, current screening methods such as NGS often require complex analysis methods that are difficult to interpret and may not accurately resolve *SMN1* copy number variants. Targeted copy number methods such as high-resolution melt (HRM) and multiplex ligation-dependent probe amplification (MLPA) are not designed to detect many pathogenic variants (18, 6). Recently, long-read sequencing has emerged as an option to identify copy number and pathogenic variants in a single workflow (19; 20). Recent advancements in sequencing technologies may help overcome these challenges by incorporating much longer unique reads that can differentiate *SMN1* from *SMN2* via paralog-specific variants. However, such assays are limited by accessibility, capital costs, and costly and complex operational infrastructure.

To address these limitations, we developed a complete assay and workflow for *SMN1* and *SMN2* genetic analysis using targeted PCR amplification, Oxford Nanopore (ONT) any-length sequencing, and a customized analytical algorithm that resolves multiple variant classes. We report results with more than 750 samples demonstrating high accuracy across diverse and complex genotypes. We also discuss operational and real-world advantages for carrier screening and diagnostic applications, including simple library preparation, flexible sample throughput, low capital investment, small instrument footprint, and modest consumable cost per sample.

## Methods

### Assay Prototyping cohort

The prototype assay was developed using genomic DNA (gDNA) from cell lines (N=97) obtained from the Centers for Disease Control and Prevention Repository (N=4), National Human Genome Research Institute (NHGRI) Sample Repository for Human Genetic Research (N=12), and National Institute of General Medical Sciences (NIGMS) Human Genetic Cell Repository (N=73) at the Coriell Institute for Medical Research (Camden, NJ; **Supplemental Table 1**). Additionally, cell lines (N=8; described in <u>21</u>) were procured from ATCC and isolated using a precipitation-based method (Qiagen, Hilden, Germany).

### Assay Optimization cohort

Human-derived, presumed normal, de-identified residual whole blood specimens (N=227) were obtained from We Are Blood (Austin, TX) under the required regulatory approvals for evaluating clinical specimens. Whole blood was purified using either silica resin/column-based method (Qiagen, Hilden, Germany) or functionalized magnetic bead (Applied Biosystems, Waltham, MA). Genomic DNA quantity (ng/uL) and quality (A_260_/A_280_) were assessed using spectrophotometry. Samples were diluted in nuclease-free water to the target concentration for analysis using the assay.

### Assay Evaluation cohort

De-identified samples with known SMA copy number status as determined by clinical testing using a droplet digital PCR assay (see below) at the CAP-accredited genomics center, at Al Jalila Children’s Specialty Hospital (Dubai, United Arab Emirates), were used for assay evaluation and test performance characterization. This study was approved by the Dubai Health Authority Research Ethics Committee (DSREC-07/2023_06 and DSREC-SR-03/2023_08).

### Droplet digital PCR

Genomic DNA extracted from peripheral blood (N = 70) were tested for *SMN1* and *SMN2* copy number status using a proprietary droplet digital PCR (ddPCR) assay (Bio-Rad, USA) following manufacturer’s instructions. This assay was clinically validated by the Al Jalila Children’s genomics laboratory (22).

### PCR and capillary electrophoresis (PCR/CE)

Only samples determined to have a homozygous deletion of the *SMN1* gene by ddPCR (N = 32) were clinically tested for *SMN2* copy number status by ddPCR. For the remaining samples with ≥ 1 *SMN1* copies (N = 44), *SMN2* dosage status was determined by the AmplideX® PCR/CE *SMN1/2* Plus Kit as previously described (23). In addition, this method was utilized to assess both *SMN1* and *SMN2* copy number for all cell line and presumed normal whole blood samples during assay development and optimization.

### PCR and Nanopore Sequencing (PCR/Nanopore)

The prototype assay workflow includes PCR master mix setup, gene-specific amplification, sample specific barcoding, paramagnetic bead size selection and concentration, library adapter ligation, Nanopore sequencing, and assay-specific analysis pipelines (**Figure 1**). Roughly 40ng gDNA was amplified by multiplex PCR targeting 2.7kb regions of *SMN1* or *SMN2* exons 7-8 and copy number neutral endogenous control amplicons from *CFTR* in one mix or a larger 11.2kb amplicon encompassing exons 3-8 in a separate mix. Reactions were cleaned up using 0.6X AMPure XP bead ratio (Beckman Coulter, Indianapolis, IN). Samples were tagged with unique barcodes during a second PCR reaction, normalized by mass after Qubit quantitation (Invitrogen, Carlsbad CA), pooled into a single reaction, and concentrated using 0.6X AMPure XP bead ratio.

**Figure 1.**
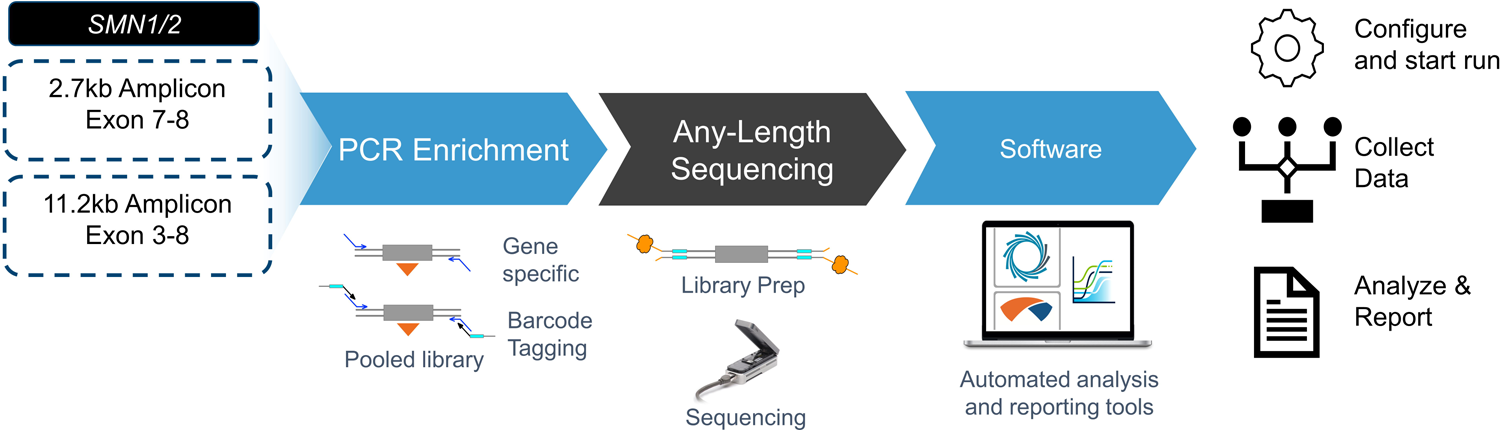
The prototype PCR/Nanopore Assay design and workflow involves two separate PCR reactions that amplify either 2.7kb or 11.2kb amplicon products. Amplicons are barcode tagged for each sample in a second PCR reaction, then pooled by mass into a single sequencing library. The library is prepared by appending nanopore-specific adaptors and loaded into a MinION flowcell for sequencing on a Mk1B connected to a computer running MinKNOW software (Oxford Nanopore Technologies). Data are analyzed with off-the-shelf and assay-specific software.

Sequencing libraries were prepared using the Nanopore Ligation Sequencing Kit (LSK-110 or LSK-114; ONT). Sequencing was conducted using MinKNOW software (22.10.10) on MinION flow cells (R9.4.1 or R10.4.1) with a Mk1B sequencer connected to a computer. Sequencing proceeded for at least 5 hours to obtain a minimum of 150 fully-spanning reads per copy per region. Super-accurate base calling was performed using Guppy (6.3.9) after sequencing and fully spanning reads were aligned to GRCh38 using minimap2 (2.15+dfsg-1).

To determine copy number, we trained a gradient boosting tree model to predict copy number using the ratio of Cs and Ts at the paralog specific variant c.840C>T in exon 7 (NM_000344.3c.840C>T; Single Nucleotide Polymorphism database, https://www.ncbi.nlm.nih.gov/snp, accession number rs1164325688, build 155, last accessed August 15^th^, 2023) (**Supplemental Figure 1**). The model incorporates read depth of fully spanning 2.7kb fragments aligned to *SMN1* (c.840C) or *SMN2* (c.840T) and the geometric mean of endogenous control amplicons (*CFTR*) to infer fold change based on normalized read depth in 2-copy calibrator samples (**Supplemental Figure 2**). The machine learning algorithm was trained on a subset of cell lines (N=79) and an independent set of residual whole blood samples (N=352). Hyperparameters for the decision tree model were selected using an 80:20 train:test split in a stratified randomly selected five-fold cross validation scheme.

Additionally, *SMN1* and *SMN2* copy number was informed with the longer 11.2kb amplicon flagged as edge cases where the machine learning algorithm predicted a copy number of 2.25 to 2.75. First, unique haplotype groups were identified and differentiated by sequence variability (e.g. SNVs) corresponding to unique copies aligning to the same region of the genome (**Supplemental Figure 3**). This allows for the identification of all unique copies for a gene target, and thus can be used to inform copy number calls and provide proper input to variant calling software that often assumes no more than two copies of the gene are present. Group sizes were normalized according to read depth and copy number was inferred for each group. The number of normalized haplotype groups were reported as the predicted copy number for samples that were flagged as edge cases when more than one group was identified.

Predicted copy number from the PCR/Nanopore assay was compared to data collected by orthogonal methods (ddPCR and/or PCR/CE). Small nucleotide variants and insertion/deletions were identified using Clair3 (24).

## Results

### Assay Development and Optimization

We developed a complementary, two-in-one MinION-based assay utilizing amplicons ranging more than 10-fold in length, including reference loci, to identify *SMN1* and *SMN2* variants. Assay designs and analysis methods were optimized for copy number changes and phased SNVs and indels (**Figure 1**). Primers were developed to amplify ∼800-3000 base fragments (“short” amplicons) encompassing both *SMN1* and *SMN2* exon 7-8 and endogenous controls (**Supplemental Figure 2**). The assay assesses *SMN1* or *SMN2* copy number by aligning reads spanning exons 7 and 8 to either *SMN1* or *SMN2*, then determining the raw read depth and ratio of reads associated with c.840C (*SMN1*) or c.840T (*SMN2*). A machine learning method infers copy number by comparing the C:T ratio in exon 7 and corresponding read depths to endogenous control amplicons in calibrator samples.

We also designed primers to amplify an 11.2kb fragment (“long” amplicon) encompassing exons 3-8 to enhance phasing SNVs in *SMN1* or *SMN2* (**Supplemental Figure 3**). This long amplicon typically captures several paralog-specific variants and facilitates sequence deconvolution of reads into haplotype groups to infer copy number. In some samples, however, there are no copy distinguishing sequence variants within one or both paralogs to independently resolve copy number.

The assay was developed and evaluated in three phases (**Figure 2**). In the prototyping phase, gene-specific primers, protocols, and reagents were designed and tested on a diverse set of cell line (N=79) and presumed normal whole blood (N=352) samples. This set included independently analyzed *SMN1* and *SMN2* genotypes ranging from 0 to ≥4 copies along with SNVs and Indels described below. Sequencing data were collected with short and long amplicons. Algorithms were trained using amplicon sequence and read depth within *SMN1* and *SMN2* compared to known genotypes for each sample. We also identified endogenous control amplicons and calibrator samples to convert *SMN1* and *SMN2* read-depth ratios into copy numbers.

**Figure 2.**
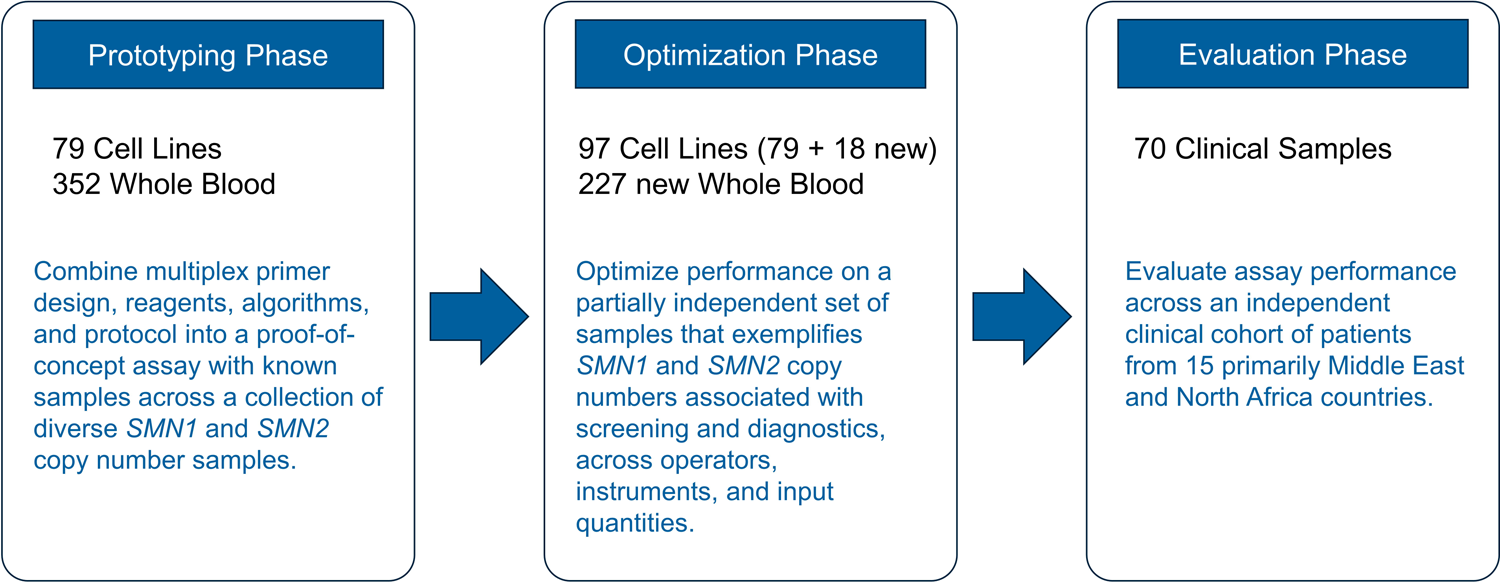
Study design for assay development consisted of three phases.

In the assay optimization phase, performance was first evaluated with the short, 2.7kb amplicon design utilizing a sample cohort of 18 additional cell lines (N=97 total) and an independent set of whole-blood samples (N=227) across 0 to ≥3 *SMN1* and *SMN2* copies (**Figure** 3**3**). Assay copy number predictions were compared to PCR/CE data and segregated by sample type.

**Figure 3.**
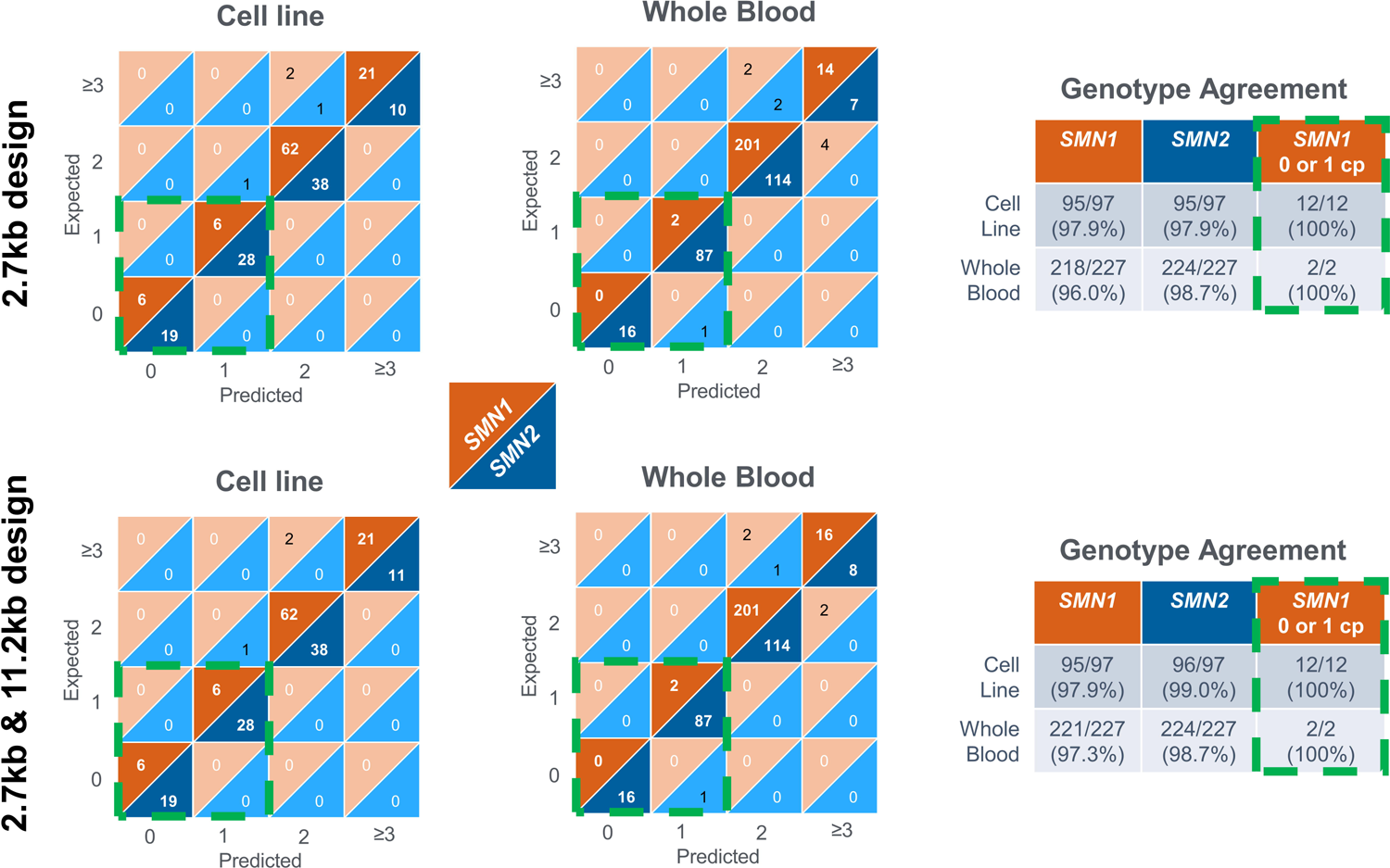
Performance of the assay to identify copy number across cell lines or presumed normal whole blood samples within the optimization data set for SMN1 (orange) or SMN2 (blue) utilizing the 2.7kb fragment or both designs where call threshold flagging would prioritize the copy number call from the 11.2kb amplicon. Genotype agreement was >97% for both SMN1 and SMN2 using both designs, and 100% for the limited number of 0 and 1 copy SMN1 samples in this data set.

Copy number agreement was 97.9% (95/97) between PCR/Nanopore and comparator method for both *SMN1* and *SMN2* in the cell-line set. One sample (NA20232) was discordant for both *SMN1* and *SMN2*. This cell line was expected to be 3 copy *SMN1* and 2 copy *SMN2* (3/2) by PCR/CE yet reported as 2/1 by the PCR/Nanopore assay. Two other samples had either *SMN1* or *SMN2* discordant copy number data where a 3 copy was expected, yet a 2 copy was predicted by the algorithm. In each case, haplotype deconvolution with the long amplicon supported the expected copy number.

In the whole blood sample cohort, 218/227 (96.0%) *SMN1* and 224/227 (98.7%) *SMN2* copy number calls agreed between PCR/Nanopore and comparator method. Of the 9 discordant samples for *SMN1*, 7 discordant samples had more than one haplotype. We used the long amplicon design to investigate these discordances, consistent with methods described. Six of the seven samples agreed with comparator data when both amplicon designs were utilized. Four of 9 discordant samples were flagged as edge cases at or near the call threshold with the short amplicon and 3 were resolved by haplotype deconvolution with the long amplicon design.

Consequently, we applied the flagging QC across the entire dataset to determine the effect on accuracy (**Figure** 3**3**). One cell line and 12 whole blood samples were flagged as edge cases at or near the call threshold with the 2.7kb amplicon for *SMN1*. Of these, 4 were discordant using the short amplicon alone, but 3 were corrected by calls using the long amplicon. Similarly, of the 3 *SMN2* calls flagged only 1 was discordant and long amplicon design supported the expected copy number. In no instance was a sample flagged where the long amplicon call disagreed with comparator data when at least 2 haplotype groups were identified. As a result, both amplicon designs were used for all subsequent analyses where call threshold flagging prioritized the long amplicon copy number call.

In addition to copy number evaluation, the assay was designed to identify and phase SNVs and indels without reflexing to other assays. For example, the *SMN2*, NM_017411.3: c.859G>C variant (dbSNP, rs121909192, build 155, last accessed August 15^th^ 2023) is associated with a less severe SMA phenotype (25, 26, 27, 28). Two variants linked with the *SMN1* duplication haplotype have been shown to flag silent carriers and increase carrier detection rates when 2 *SMN1* copies occur on the same chromosome. These variants are NM_000344.3:c.*3+80T>G (alias g.27134T>G; dbSNP, rs143838139, build 155, last accessed August 15^th^ 2023) and NM_000344.3: c.*211_*212del (alias g.27706_27707delAT; dbSNP, rs200800214, build 155, last accessed August 15^th^ 2023).

PCR/Nanopore genotyping of these variants agreed with the PCR/CE comparator method in 324/324 samples for c.*3+80T>G and c.859G>C, and in 322/324 (99.4%) for c.*211_212del (**Figure 4**). Investigation of the two discordant samples suggested erroneous comparator data. In HG00691, a rare non-pathogenic deletion in *SMN2* (rs576032516) shifted a CE peak into the mutant CE bin, resulting in a false positive. In the second sample, the automated peak caller in the CE assay incorrectly called noise along the baseline within the c.*211_212del bin, resulting in another false positive call.

**Figure 4.**
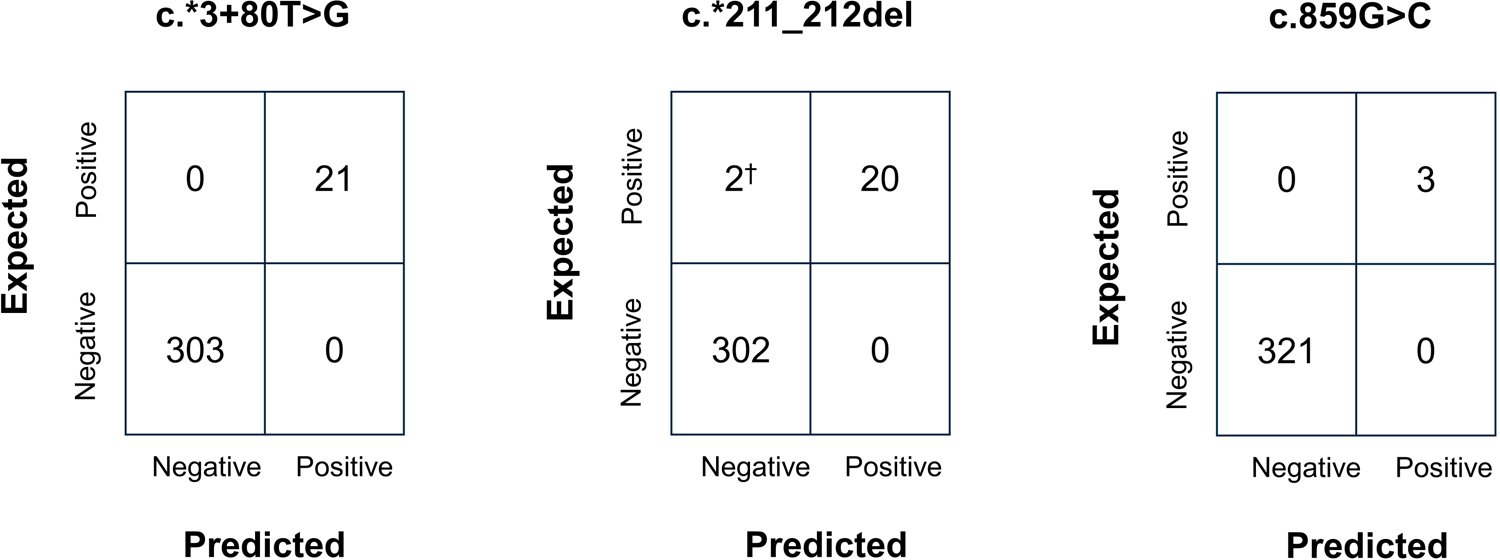
Confusion matrix comparing the expected and predicted SNVs associated with silent carrier or disease modifier risk. The algorithm accurately identified each variant type except for two. †Inspection of the underlying trace data for these two samples in the comparator assay revealed a profile consistent with false-positive calls and a defined root cause in each case. This correction would result in 100% agreement across each variant for the PCR/Nanopore assay.

### Independent Evaluation of Residual Clinical Samples

Next, we evaluated assay performance across a diverse, independent cohort of samples from 70 individuals (57% females, average age 2.17 years, range 1 day – 16 years) with known *SMN1* and *SMN2* copy number status originally tested at Al Jalila Children’s Specialty Hospital, Dubai, UAE. Those individuals represented 14 countries, primarily from the Middle East and North Africa (**Supplemental Table 2**).

Of all individuals, 30 (42.9%) had homozygous deletions in the *SMN1* gene, while the remaining 40 (57.1%) had 1 to 4 copies of this gene. Sixty-seven of the 70 samples were also evaluated for *SMN2* copy number. Most individuals (58%) had 2 *SMN2* copies, 10% had 0 copies and 42% had 1, 3, or ≥ 4 copies of this gene (Supplemental Table 3).

The PCR/Nanopore assay correctly identified 70/70 and 65/67 of *SMN1* and *SMN2* copy number states for a concordance rate of 100% (95% CI, 94.80% - 100.00 %) and 97.0% (95% CI, 89.75% - 99.18%), respectively (**Table 1**).

**Table 1.** PCR/Nanopore assay performance based on concordance data using clinical samples with known SMN1/2 copy number status.

| COPY NUMBER | NUMBER OF CLINICAL BLOOD SAMPLES (BOTH DESIGNS) |
| --- | --- |

| STATE | SMN1 (N=70) | SMN2 (N = 67) |
| --- | --- | --- |
| 0 | 30/30 | 7/7 |
| 1 | 8/8 | 7/7 |
| 2 | 22/22 | 37/39 |
| ≥ 3 | 10/10 | 14/14 |

### Cost Effectiveness and Hand-On Time

The accessibility and broad use of an *SMN1/2* screening and genotyping assay depends on numerous criteria, including performance, operational factors and cost. Assay workflow, analysis and economic considerations are especially important for laboratories in lower resource environments. We assessed these elements by performing a time-motion analysis, quantifying the scalability of the assay across different sample batch volumes, and calculating estimated costs per sample.

Time-motion analysis was estimated across 6 operators based on experience for both a 24- and 96-sample batch (**Supplemental Table 4**). The workflow from sample to answer requires less than 48 hours for a 24-sample batch on a Mk1B connected to a computer with a recommended GPU. Gene-specific and barcoding PCR can be completed in a single work shift. Subsequently, samples can be pooled, the library prepared, and sequencing initiated with sequencing and analysis completed overnight for review the next day. A 96-sample batch required additional sequencing and data processing time but was still completed within 72 hours.

The assay supports 12 to 96 samples per batch (**Supplemental Table 5**). Batch size is currently only limited by the recommended barcodes available from ONT (N=96) since flow cells can routinely process >10M reads. A single Mk1B running 12 samples once per week could screen 624 samples per year. By comparison, 480 samples could be processed per batch on one GridION with 5 independently accessible flow cells, real-time base calling, demultiplexing, and alignment. Utilizing 3 staggered run batches per week a lab could process 75,000 samples per year. This throughput can scale linearly by adding additional GridION instruments. In addition, the assay supports automated reaction setup, bead isolation, quantitation, and data analysis to further improve workflow efficiency.

We estimated less than $20 USD per sample for materials costs excluding PCR (**Supplemental Table 6**). Since the assay is still in development, PCR enrichment costs cannot be accurately determined. For example, the assay design must be finalized and verified, reagents and kits manufactured, and quality control and release testing established. However, PCR is well documented to be highly cost-effective at scale, and we expect that the total per-sample costs will be comparable to other *SMN1/2* diagnostic kits that provide far less genotyping information and insight. These costs also do not include instrument-related expenses. Importantly, ONT instruments (e.g. Mk1B and GridION) have low capital requirements and benchtop footprints, which creates flexibility for laboratories in how they implement the technology. For either instrument, a computer is required for data analysis. For Mk1B instruments, the computer must be equipped with a high-performance GPU to utilize live basecalling, demultiplexing, and alignment during sequencing.

## Discussion

We developed a novel dual-mode PCR/Nanopore sequencing assay for comprehensive, scalable and cost-effective *SMN1/2* genotyping. The assay design utilizes a PCR target enrichment approach to generate 2.7kb “short” and 11.2kb “long” amplicons spanning regions of the *SMN1* and *SMN2* genes, as well as a machine learning-based analytical pipeline haplotype phasing and read depth data to decipher sequence and copy number variants specific to both highly homologous genes.

Data from both amplicons were combined to achieve the highest accuracy for the prototype assay (>97%). Other long-read sequencing designs utilize only phased haplotype analysis either by hybrid capture (20) or multiple ultra-long (>26kb) amplicons on the PacBio SMRT platform (19). Robustness may be adversely affected by identical haplotypes without read depth normalized to endogenous and exogenous control amplicons. Indeed, we observed identical haplotype groups in our sample set when only the long amplicon was used for copy number evaluation. In contrast, the PCR/Nanopore assay utilizes a combination of read-depth normalization (short amplicon) and haplotype phasing (long amplicon) to resolve *SMN1* and *SMN2* copy number and phase pathogenic SNV/introns. In addition, the assay scales from tens to tens of thousands of samples per year with reduced capital and per-sample costs compared to other sequencing-based assays. Reducing costs is critical for many existing screening labs or those looking to adopt such screening assays, as reimbursement costs are not always able to cover test costs with NGS workflows. Lastly, the assay includes analysis and reporting software to reduce interpretation expertise and overhead, though the full suite of push-button automated analysis software is still in development.

We acknowledge a few limitations of the current study. The optimized prototype was designed to phase variants across *SMN1* or *SMN2* exons 3-8. Although we demonstrated variant phasing with non-pathogenic silent carrier and disease modifier SNVs, full exon coverage of *SMN1* and *SMN2* is preferred to assess all potential SNV/indel variants. In ongoing work, we have extended the assay design to cover exons 1, 2a, and 2b, though performance has not yet been evaluated. Further, amplification efficiency must be optimized to reduce read depth variability between samples by optimizing primers and cycling conditions. These modifications are expected to improve the resolution and differentiation of 3- and 4-copy genotypes, especially important for treatment decisions that may rely on accurate high copy number *SMN2* calls when *SMN1* is not detected. We have also begun evaluating additional endogenous control amplicons to improve accuracy. Finally, we recognized that amplicon-based methods can be affected by sample-specific SNVs in primer-binding regions. This risk was reduced by utilizing two different primer sets across different amplicon sizes to resolve copy number.

In summary, we present results from the development, optimization, and external evaluation of a novel PCR/Nanopore assay using over 750 samples, including cell lines, residual presumed normal blood donors, and patient specimens with known *SMN1* and *SMN2* genotypes. The results reveal accurate detection of multiple categories of clinically informative variants, including SNVs, indels and CNVs. SNV phasing was demonstrated through known silent carrier (2+0) and disease-modifier variants. Importantly, the assay is cost-effective and scalable, showing potential for broad implementation in diagnostic and screening programs. Last, we note a key benefit of the assay chemistry, platform and workflow is its flexibility to include additional, variants in *CFTR*, *FMR1*, and *HBA1/2* and other genes associated with commonly screened genetic disorders. This extensibility may further expand the utility of the approach and application and represents an important future direction for technology development.

## Abbreviations

SMA: Spinal Muscular Atrophy

SNVs: Single nucleotide variants Indels, Insertions and deletions

CNVs: Copy number variants

NGS: Next generation sequencing

US: United States

FDA: Food and Drug Administration

HRM: High-resolution melt

MLPA: Multiplex ligation-dependent probe amplification

ONT: Oxford Nanopore Technologies

PCR: Polymerase chain reaction

gDNA: Genomic DNA

NHGRI: National Human Genome Research Institute

NIGMS: National Institute of General Medical Sciences

ddPCR: droplet digital PCR

CE: Capillary electrophoresis

QC: Quality control

## Data Availability

All data produced in the present work are contained in the manuscript

## Acknowledgements

We would like to thank all members of Al Jalila Children’s Genomics Center, Asuragen, and Oxford Nanopore Technologies, specifically Frédérique Lerêteux, Rita Aoun, and Hannah Lucio, for their valuable input on this work.

