## Supplementary figures and images for "A dual-mode targeted Nanopore sequencing assay for comprehensive *SMN1* and *SMN2* variant analysis"

### Supplemental Figure 1

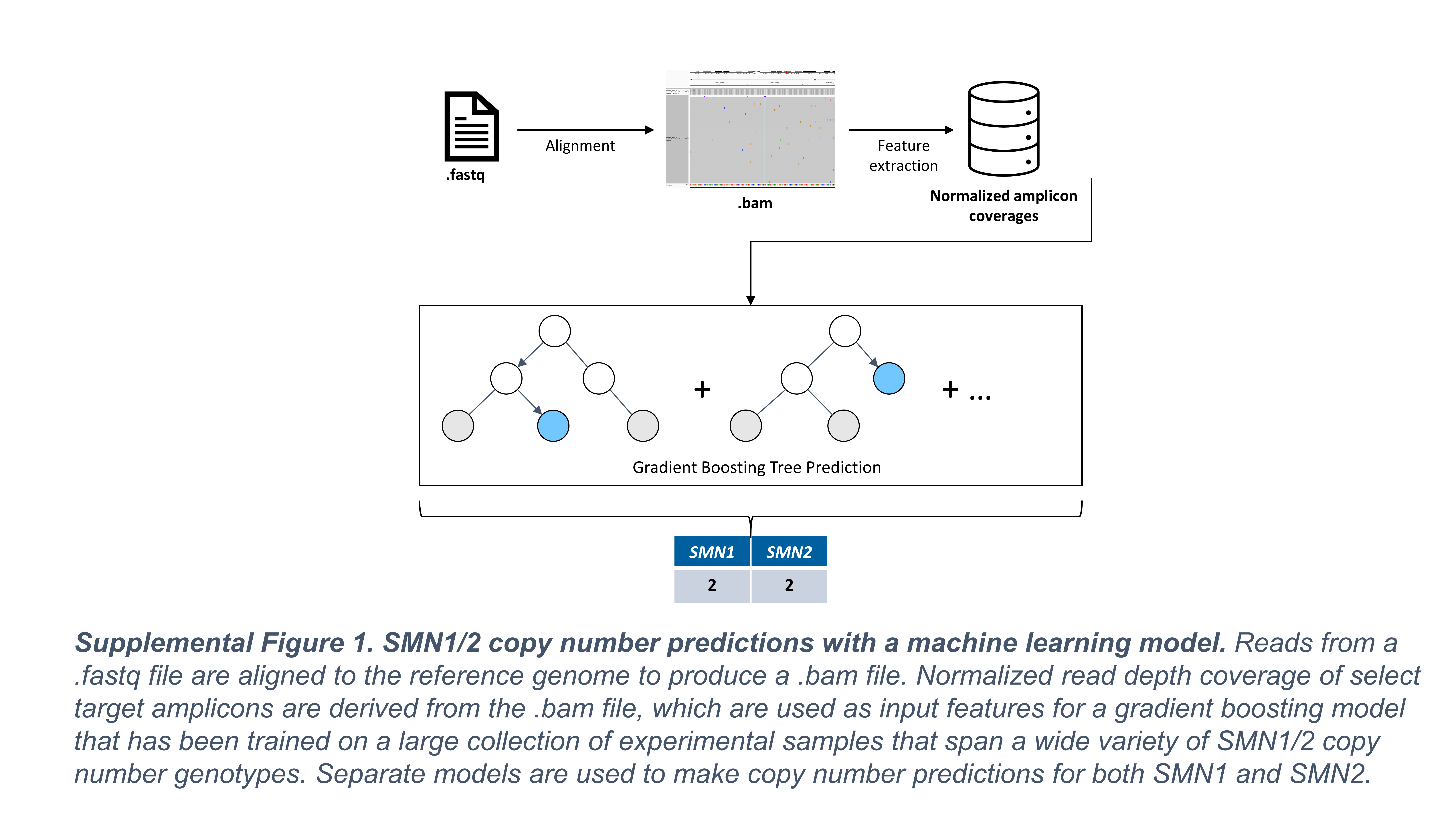

### Supplemental Figure 2

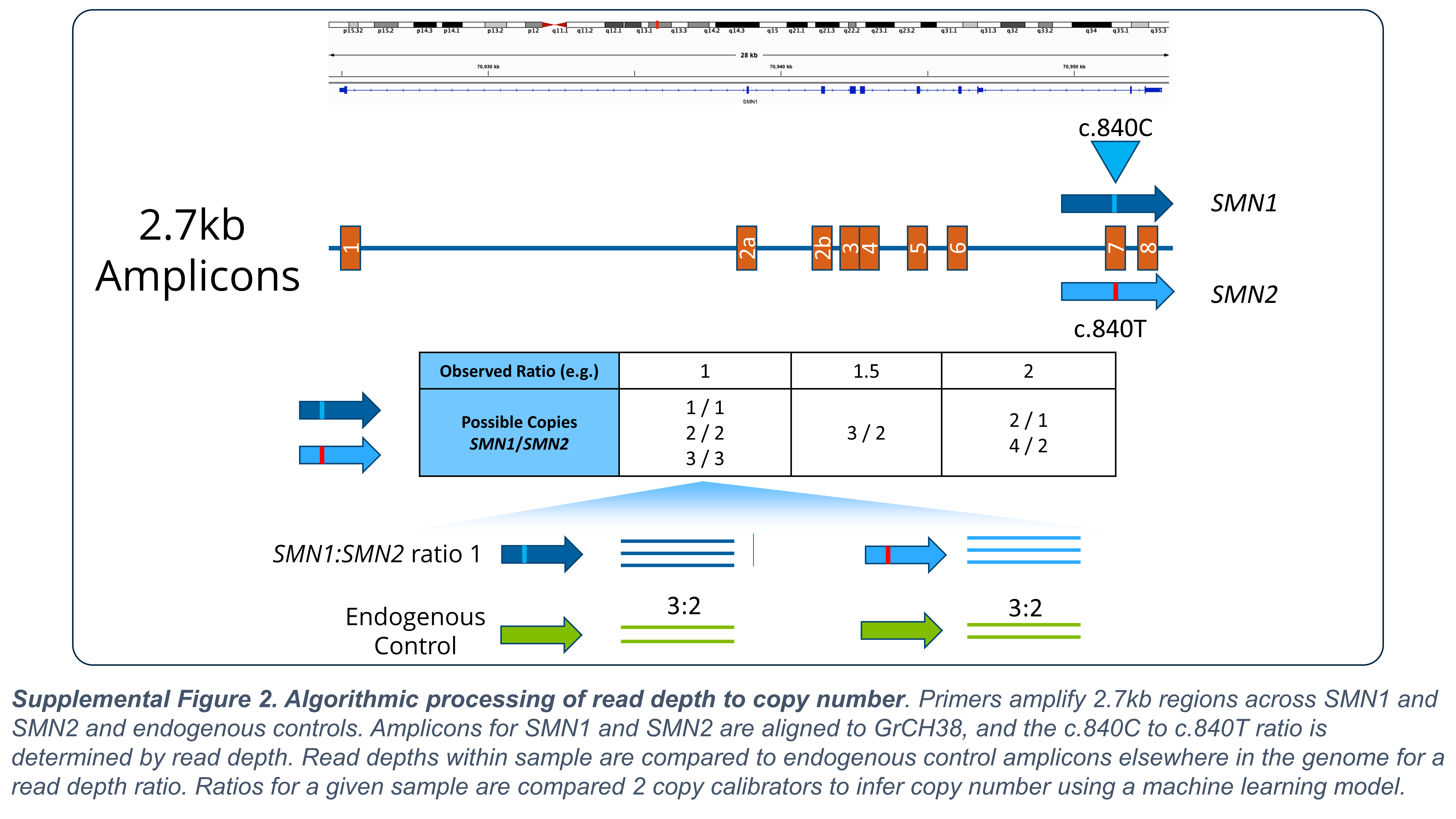

### Supplemental Figure 3

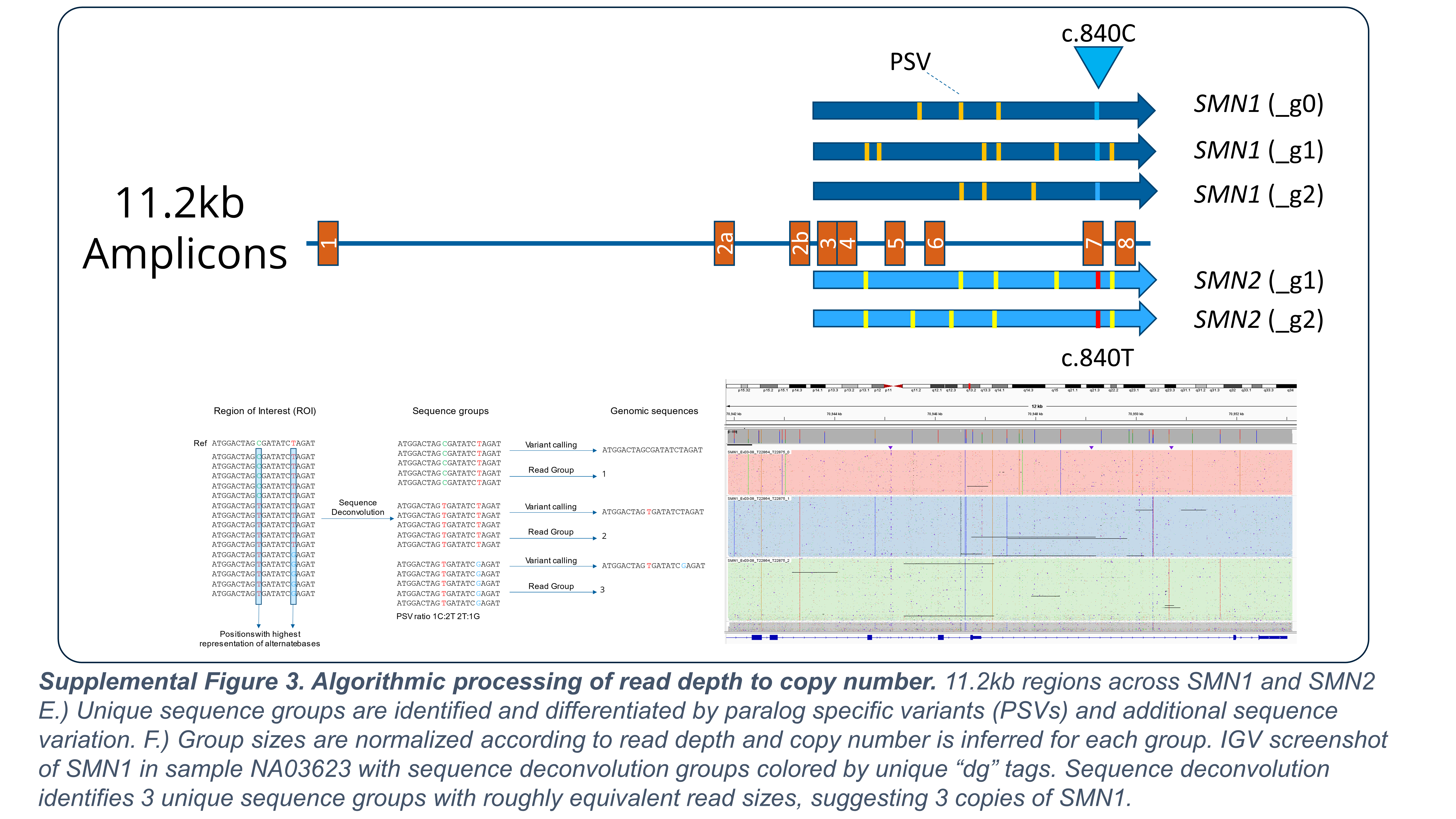
